# Integrative proteogenomics study identifies the indirect effect of circulating proteins on 8 diseases via telomere length

**DOI:** 10.1101/2023.12.06.23299603

**Authors:** Shifang Li, Meijiao Gong

## Abstract

Growing evidence has revealed the associations between telomere length and diseases; however, the mechanisms behind these links are not fully understood. In this study, by applying proteome-wide Mendelian randomisation (MR), multiple sensitivity analyses, colocalization, single-cell RNA-sequencing, and mediation analysis, the potential mechanisms that link 4,907 plasma proteins, telomere length, and 8 diseases were identified. Following MR analysis for the effects of plasma protein levels on telomere length, 34 proteins were found to be causally related to telomere length. Among these, 8 proteins (PSMB4, PARP1, GDI2, MAX, GMPR2, ARPC1B, ATOX1, and NUDT5) were found to be well colocalized with telomere length (posterior probability >80%). 21 mediation pairs were revealed for the indirect effect of circulating proteins on 8 diseases via telomere length. Furthermore, 35 proteins, including CD8A, were found to be influenced by telomere length among 4,907 proteins. No significant mediation pairs for circulating proteins that mediate the effects of telomere length on disease have been identified. Overall, our study provided insights into understanding the biology of telomeres and prioritized the identified proteins as prospective intervention targets for the disease.

## Introduction

Telomeres are 6-nucleotide repeats that are found at the ends of chromosomes and are normally reduced as cells replicate and age. When telomeres reach a critical length, cells enter senescence; thus, telomere shortening and malfunction are one of the major characteristics of aging [1-2]. At the population level, there is significant individual variation in mean telomere length (TL), which is typically assessed in leukocytes but also in other tissues (eg., testis and cerebellum) [2-3]. Based on the findings of epidemiological studies and Mendelian randomisation (MR) analyses, researchers found out that several diseases, including melanoma, prostate cancer, thyroid, uterine fibroid, multiple sclerosis, idiopathic pulmonary fibrosis (IPF), coronary artery disease, and hypothyroidism are associated with telomere dysfunction; however, the underlying mechanism between these associations is not fully understood [3-8]. Identifying circulating proteins that mediate these associations or determining the indirect influence of circulating proteins on diseases via TL is an approach for disentangling this association. As circulating proteins in the blood may be evaluated and, in some cases, manipulated, identifying related proteins may provide insights into developing probable therapeutic targets [9].

MR is a classical approach in genetic epidemiology research that employs single nucleotide polymorphisms (SNPs) related to the exposure variable under inquiry. Using genetic polymorphisms randomly assigned at conception as an instrumental variable (IV) for exposure, it is feasible to obtain estimates that are less susceptible to the influence of environmental confounders and reverse causation [10-11]. Researchers are currently using this strategy to uncover therapies for various diseases, including stroke, idiopathic pulmonary fibrosis (IPF), COVID-19, and ischemic heart disease [9,12-14]. In addition, by performing MR and mediation analysis, Yoshiji *et al*. have discovered nephronectin as an actionable mediator of the effect of body mass index (BMI) on COVID-19 and COL6A3 as a mediator of the effect of obesity on coronary artery disease [9,15]. Similarly, Chen *et al*. demonstrated that BMI, white matter hyperintensity, and atrial fibrillation appear to mediate the TFPI, IL6RA, and TMPRSS5 correlations with stroke [13]. The findings from these studies suggest that an MR-based strategy represents an intriguing strategy for illuminating the potential mechanisms between risk factors and disease.

In this study, we utilized proteome-wide MR using large-scale GWASs of plasma protein, multiple sensitivity analyses, colocalization, single-cell RNA-sequencing (scRNA-seq) analysis, and mediation analysis to identify the indirect effect of circulating proteins on 8 diseases via TL and uncover circulating proteins mediating the effects of TL on disease.

## Methods

### Data source for MR analysis

For MR analysis, publicly available GWAS summary data were applied. We selected the most complete GWAS for plasma proteins, which includes genome-wide associations of genetic variants for 4,907 circulating proteins in 35,559 Icelanders [16]. Prostate cancer (access ID: GCST90274714), uterine fibroids (access ID: GCST009158), hypothyroidism (access ID: GCST90204167), coronary artery disease (access ID: GCST005194), melanoma (access ID: GCST90011809), and thyroid cancer (access ID: GCST90011813) summary statistics were retrieved from the GWAS Catalog (https://www.ebi.ac.uk/gwas/). Prostate cancer included 122,188 cases and 604,640 controls of European ancestry [17], uterine fibroids comprised 20,406 cases and 223,918 controls of European ancestry [18], hypothyroidism included 51,194 cases and 443,383 controls of European ancestry [19], coronary artery disease comprised 34,541 cases and 261,984 controls of European ancestry [20], melanoma included 6,777 cases and 410,350 controls of European ancestry [21], and thyroid cancer included 762 cases and 410,350 controls of European ancestry [21]. The GWAS summary statistics for multiple sclerosis (access ID: ieu-b-18) [22], TL (access ID: ieu-b-4879) [8], and 102 CD8+ T cells subpopulations [23] (**Supplementary Table 1**) were downloaded from the ieu open gwas project (https://gwas.mrcieu.ac.uk/datasets/). The summary statistics of the large GWAS of IPF were applied from https://github.com/genomicsITER/PFgenetics [24]. Details of the included GWASs are summarized in **Supplementary Table 1**.

### IVs selection

To satisfy the MR assumptions, the SNPs chosen as IVs had to show strong associations with exposure. To identify SNPs strongly linked to all symptoms, a genome-wide significance criterion of *p*<5e-08 was utilized. SNPs in the MHC region were removed as well in the case of the complex linkage disequilibrium (LD) structure of the MHC locus. To address bias caused by LD, we implemented SNP clumping with parameters kb=10,000 and r^2^=0.001, as described previously [25]. Furthermore, we removed palindrome SNPs and used the F-statistic to measure the strength of each SNP. SNPs with F-statistics greater than 10 were deemed sufficiently powerful to serve as IVs for the trait.

### MR and colocalization analysis

The MR analysis was carried out as previously described [25]. In brief, the inverse-variance weighting approach (IVs≥2) and Wald ratio (IV=1) were chosen as the primary analysis methods in the present study. Sensitivity analyses were conducted to ensure the robustness of the findings and to resolve any potential breaches of the MR assumptions. Cochran’s Q test was employed to evaluate SNP heterogeneity in both the IVW and MR Egger methods. In a heterogeneity test, I^2^ statistics were estimated using the “Isq()” function and heterogeneity *p*-value with the “mr_heterogeneity()” function. Following that, a “leave one out” analysis was executed to assess the impact of particular SNPs on the assessment of causal effects. All MR analysis were performed in R version 4.2.2 using the “TwoSampleMR” package [26]. The Benjamini-Hochberg multiple testing was used to account for multiple tests. Colocalization analysis was used to assess the possibility of two traits sharing a shared causal variant, as previously stated [25, 27]. A posterior probability for H4 (PH4) of more than 80% was considered significant colocalization evidence.

### Mediation analysis

To evaluate the causal mediation effects, network MR with a product of coefficients method was employed, as described by Yoshiji and others [9, 15]. In brief, we estimated the effect of plasma protein levels on TL (β_protein-to-TL_) and the effect of TL on diseases (β_TL-to-disease_), and then multiplied these values (β_mediated_=β_protein-to-TL_×β_TL-to-disease_). Subsequently, we divided β_mediated_ by β_total_ (β_protein-to-disease_) to estimate the proportion mediated.

### Single-cell RNA sequencing analysis

The GSE138266 [28] and GSE174072 [29] datasets were obtained from the NCBI Gene Expression Omnibus (GEO) and analyzed using the R software. GSE138266 includes 12 scRNA-seq datasets from cerebrospinal fluid (CSF) in 6 multiple sclerosis patients and 6 control groups. The raw data of the gene expression matrix was taken from GEO and turned into a Seurat object for downstream processing, as previously indicated [30]. In brief, doublets were detected using the “scrub.scrub_doublets()” function in Scrublet package (v2.0.2) [31] and further removed. The Harmony R package v1.0 was used for addressing batch effects after normalizing the data with log transformation and scaling [32]. The “FindClusters” function was used to cluster cells, which were then annotated according to the original literature [28, 29]. Gene sets (PSMB4, PARP1, GDI2, MAX, GMPR2, ARPC1B, ATOX1, and NUDT5) for each cell type were used for scoring, and cell scoring was performed by the “AddModuleScore” function in Seurat (v4.2.0).

## Results and discussion

### Identification of causal plasma proteins to TL

First, we implemented two-sample MR to determine the causal effects of plasma proteins on TL outcomes. For the purpose of this analysis, *cis*-acting protein quantitative loci (*cis*-pQTLs, pQTLs that are present inside a 1 Mb region around the gene body) from 4,907 proteins were utilized, reducing the probability of directing horizontal pleiotropy [9,12,14]. Following the *cis*-pQTL search and data harmonization, 1,428 proteins were evaluated in MR for their estimated causal effects on TL (**Figure 1A**). The F-statistics for all of the examined proteins were all greater than 10, minimizing the possibility of weak instrument bias significantly (**Supplementary Table 1**). After applying the Benjamini-Hochberg multiple testing correction (FDR<0.05) and filtering procedure (**Figure 1A**), MR revealed 34 causal proteins, including 17 positive and 18 negative associations (**Figure 1B**). For example, one standard deviation (SD) increase in genetically predicted GDI2 levels was associated with decreased TL (odds ratios (OR)=0.84, 95% confidence intervals (CI) 0.80-0.87, *p*=2.08e-15). Similarly, a one standard deviation rise in genetically predicted ARPC1B levels was linked to lower levels of TL (OR=0.88, 95%CI 0.85-0.93, *p*=7.78e-07). Higher genetically predicted levels of NUDT5 (OR[95%CI]=1.12[1.06,1.18]; *p*=3.12e-05) and PARP1 (OR[95%CI]=1.88[1.72, 2.06]; *p*=1.61e-44) have been associated with increased levels of TL. The MR-Steiger test was used to investigate potential bias from reverse causation, which supported a causal direction of these 34 plasma protein levels influencing TL outcomes (**Supplemental Table 1**). The 18 proteins found to be negatively linked to TL were involved in the proteasome core complex, NIK/NF-kappaB signaling, Wnt signaling pathway, and interleukin-1-mediated signaling network (**Figure 1C**). Furthermore, 17 proteins positively related to TL were shown to be enriched in cellular response to starvation, RNA helicase activity, mitochondrial DNA metabolic process, and antifungal humoral response (*p*.adj<0.05) (**Figure 1C**). Colocalization analyses were then performed to assess the likelihood that protein and TL share the same causal variants rather than being shared accidentally due to LD associations (**Figure 1D-1E**). Interestingly, 8 proteins (PSMB4, PARP1, GDI2, MAX, GMPR2, ARPC1B, ATOX1, and NUDT5) were well colocalized with TL, with posterior probability greater than 80% for a shared signal. Two proteins (NRBP1 and BTN3A1) have a medium posterior probability of more than 70%. The other 25 proteins had a poor colocalization posterior probability with TL, indicating that these interactions may be impacted by LD.

**Figure 1.**
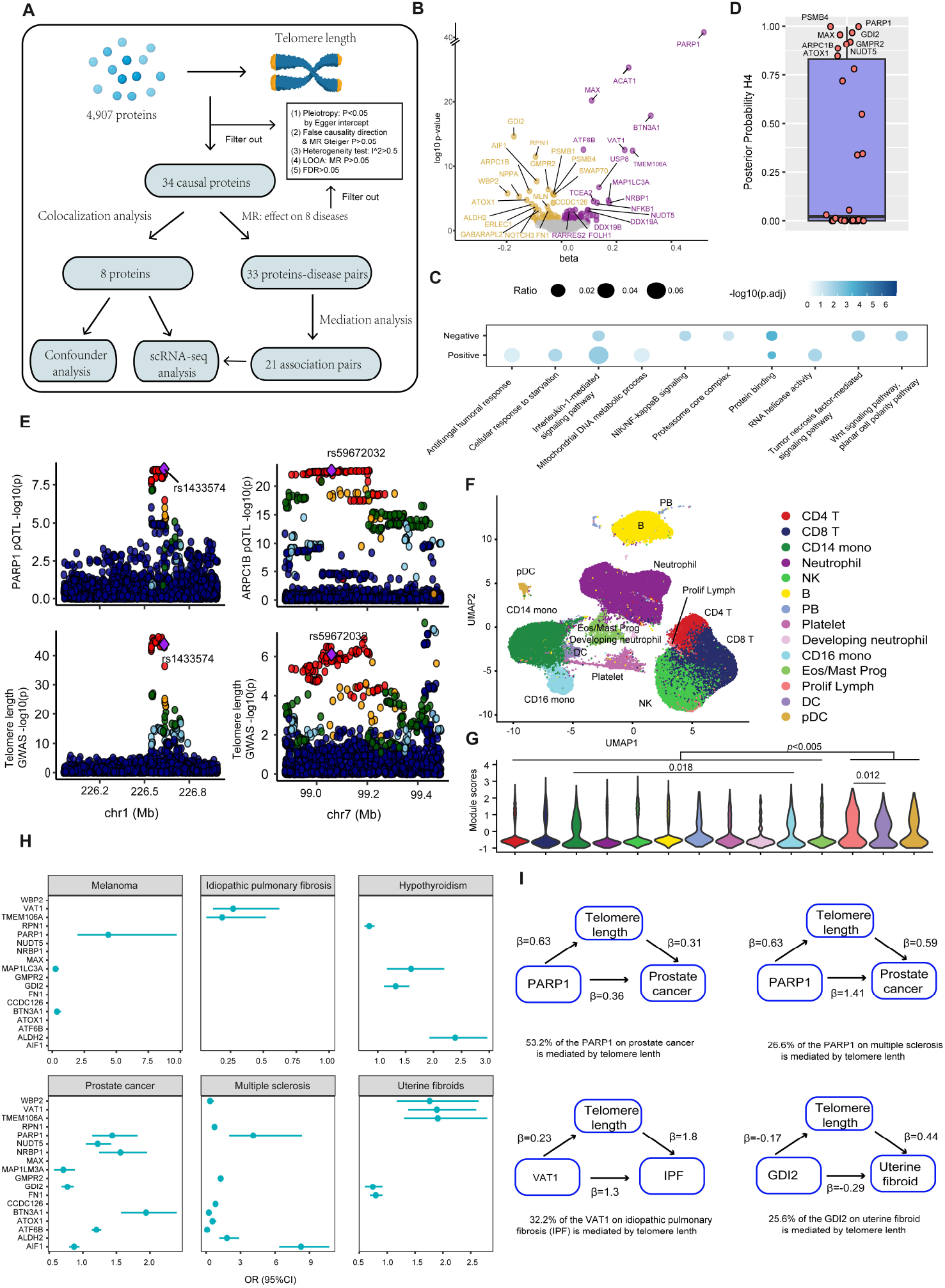
The causal effect of plasma proteins on TL. (**A**) Design overview for the investigation of the causal effect of proteins on TL. FDR, false discovery rate; LOOA, leave-one-out analysis. (**B**) Volcano plots of MR results for the causal effect of plasma proteins on TL levels. Labeled proteins are 34 proteins examined in the primary analysis with a significant FDR-corrected *p*-value<0.05. (**C**) Biological function enrichment analysis for the 17 positively and 18 negatively associated TL proteins. (**D**) A boxplot illustrating the results of the co-localization analysis for the 34 proteins described in (B). Eight proteins with high colocalization evidence (PH4>0.8) were labeled. (**E**) The locus-compare scatter plot for the identified proteins’ association with TL. The locus-compare scatter plot compares the results of the protein quantitative trait loci (pQTL) and TL genome-wide association study (GWAS), indicating whether the two traits shared the same genetic variants. (**F**) UMAP embeddings of scRNA-seq profiles (GSE174072) from 62,164 blood leukocytes in 12 healthy donors. (**G**) Module scores of gene sets (PSMB4, PARP1, GDI2, MAX, GMPR2, ARPC1B, ATOX1, and NUDT5) in each cell type. The *p*-value was obtained by the Mann-Whitney U test. (**H**) Causal effects of the selected proteins on diseases using MR analysis. The dots are the causal estimates on the OR scale, and the whiskers represent the 95% confidence intervals (CI) for these ORs. OR, odd ratios. (**I**) Mediation effects of selected proteins on diseases via TL. Mediation analyses were used to quantify the effects of 34 proteins on 8 diseases outcomes via TL. The global mediation-pairs were provided in Supplemental Table 1. TL, telomere length.

To validate the assumption of a lack of directional pleiotropy, which can reintroduce confounding, we utilized the PhenoScanner (http://www.phenoscanner.medschl.cam.ac.uk/) databases to find out whether the *cis*-pQTLs for 8 proteins were associated with any traits or diseases at the genome-wide significant threshold of *p*<5e-08. Except for MAX and GMPR2, no genome-wide significant association was detected for six proteins (PSMB4, PARP1, GDI2, ARPC1B, ATOX1, and NUDT5). The deCODE study’s lead *cis*-pQTL for MAX (rs8181938) and GMPR2 (rs34354104) were associated with body-related traits (termed as sub-associations), such as height, forced vital capacity, mean corpuscular hemoglobin, and mean corpuscular volume, among others (**Supplementary Table 1**); yet, since *cis*-acting protein quantitative loci were utilized in our study, these associations may represent a case of vertical pleiotropy, minimizing the probability of directional pleiotropy. Indeed, when employing *cis*-pQTL for MAX (rs8181938) and GMPR2 (rs34354104) from the deCODE study as exposure and these sub-association as outcomes, MR analysis indicated that both MAX and GMPR2 levels were supposed to affect these sub-associations with no reverse causation (*p*<4.2e-08, MR-Steiger test), suggesting that these sub-associations may be a case of vertical pleiotropy, which does not bias MR interpretation (**Supplemental Table 1**).

To determine whether the 8 causative proteins had any cell type-specific enrichment in whole blood leukocytes, we performed scRNA-seq analysis from GEO (GSE174072) [29], which included 62,164 cells from 12 healthy individuals (**Figure 1F**). 14 cell types were annotated based on the metadata of cells in the origin paper [29]. Subsequently, we compared module scores across cell populations to uncover a probable cell population linked to these 8 causative proteins (**Figure 1G**). The module score of proliferating lymphocytes, dendritic cells (DCs), and plasmacytoid DCs was found to be considerably higher than the other cell populations (Mann-Whitney U test, *p*<0.001), implying that these cell types may be responsible for the local production of the 8 proteins.

### Mediation effect of proteins on 8 diseases via TL

Previous study has demonstrated that TL has a causative effect on various diseases, including an increased risk of melanoma, prostate cancer, thyroid cancer, uterine fibroid, multiple sclerosis, and idiopathic pulmonary fibrosis with each standard deviation increase in TL [8]. In contrast, each standard deviation increase in TL was related to a lower risk of coronary artery disease and hypothyroidism [8]. However, the mechanisms behind these associations were primarily unresolved. To investigate this, two hypotheses have been proposed: (1) TL may participate in protein mediation of diseases; and (2) proteins mediate the influence of TL on diseases (discussed below). To test the first hypothesis, a two-sample MR was used to investigate the causal effect of 34 proteins related to TL on 8 diseases (**Supplemental Table 1**). Following MR analysis, sensitivity analysis identified 33 substantially associated associations (two associations had been removed due to failure to pass leave-one-out analysis) (**Figure 1H**). Except for BTN3A1, which appears to be shared as a causative variant with prostate cancer (PH4=0.807), there is no strong colocalization evidence for these significant associations. Interestingly, genetically higher levels of PARP1 were related to an increased incidence of melanoma (OR[95%CI]: 4.37[1.96, 9.71]; *p*=0.00029), prostate cancer (OR[95%CI]: 1.43[1.14, 1.81]; *p*=0.0021), and multiple sclerosis (OR[95%CI]: 4.09[2.009, 8.34]; *p*=0.0001).

Furthermore, ATOX1 was found to be genetically linked to a decreased risk of multiple sclerosis. Using sing-cell RNA sequencing of cerebrospinal fluid from multiple sclerosis patients and control groups (GEO, GSE138266) [28], it was found that PARP1 expression in B cells and plasma cells was higher in multiple sclerosis patients than in control groups (*p*<0.05) (**Figure S2A-S2D**). In contrast, ATOX1 expression in mDCs was lower in the cerebrospinal fluid of multiple sclerosis patients than in control groups (*p*=0.03). These findings suggest that PARP1 and ATOX1 may represent potential therapeutic targets for multiple sclerosis.

Subsequently, we performed a mediation study utilizing the effect estimates from two-step MR and the overall effect from primary MR to evaluate the indirect effect of proteins on 8 diseases via TL (**Figure 1I**). There were a total of 21 mediation pairs revealed (**Supplemental Table 1**). For example, the proportion of PARP1’s TL mediation effect on melanoma, prostate cancer, and multiple sclerosis is 19.1%, 53.2%, and 26.6%, respectively. Similarly, with values of 11%, 20.4%, and 25.6%, TL appears to mediate the effect of GDI2 on hypothyroidism, prostate cancer, and uterine fibroids, respectively. The indirect effect of VAT1 on IPF risk via TL accounts for 32% of the total. Similarly, the proportion of TMEM106A’s mediation effect on IPF via TL is approximately one-quarter of the entire effect (28%) (**Figure 1I**).

### Identification of the causal protein affected by TL

Following that, we used two-sample MR to quantify the effect of TL on circulating protein levels on a proteome-wide scale (**Figure 2A**). The F-statistic, which measures the strength of the correlation between genetic variations and TL, was 119.6, indicating that there was no evidence of instrument bias. Out of the 4,907 proteins examined (**Supplemental Table 1**), 40 were found to be influenced by TL (FDR<0.05), suggesting that TL has a moderate effect on plasma protein levels. After that, we performed heterogeneity test, directional pleiotropy test, and reverse-causation test to keep only the proteins that had been significantly affected by TL. We found no significant heterogeneity in 40 proteins (I^2^ <50% for all). The MR-Egger intercept test revealed no evidence of directed horizontal pleiotropy in 35 of the 40 proteins tested (*P*_Egger intercept_>0.05). There was no evidence of reverse causation using protein levels as the exposures and TL as the outcome. Importantly, according to the leave-one-out analysis, these 35 associations were not driven by outliers. Therefore, a total of 35 protein levels were identified as causal proteins impacted by TL, with no apparent heterogeneity, directional pleiotropy, or reverse causation (**Figure 2B-2C**). Among the 35 causal proteins, 24 positively proteins related to TL were positively associated with antimicrobial humoral response (*p*.adj=0.00035) and positive regulation of wound healing (*p*.adj=0.0038), while 11 negatively proteins associated with TL were negatively associated with cellular protein metabolic process (*p*.adj=0.018) and negative regulation of IRE1-mediated unfolded protein response (*p*.adj=0.018) (**Figure 2D**). Interestingly, among the 35 TL-driven proteins studied, higher levels of TL were shown to be related to higher levels of plasma CD8A (OR[95%CI]: 1.25[1.15, 1.36]; *p*=1.15e-07) (**Figure 2C**). Previous research has shown that the number of CD8+ T cells declines linearly with age, which is consistent with this finding [33-34]. To figure out whether the subpopulation of CD8+ T cells was influenced by TL, MR analysis was carried out on 102 cell populations of CD8 T cells from a cohort of 3,757 Sardinians [23]. Interestingly, following multiple testing corrections (FDR<0.05), CD8+ Natural Killer T cells, Naive CD8+ T cells, and CD28+ CD45RA+ CD8+ T cells were essentially affected by TL (**Figure 2E**). For these causality associations, MR analyses using the weighted median, weighted mode, and MR-Egger slope approaches produced directionally consistent results, as accomplished by the inverse variance weighted method (**Supplemental Table 1**).

**Figure 2.**
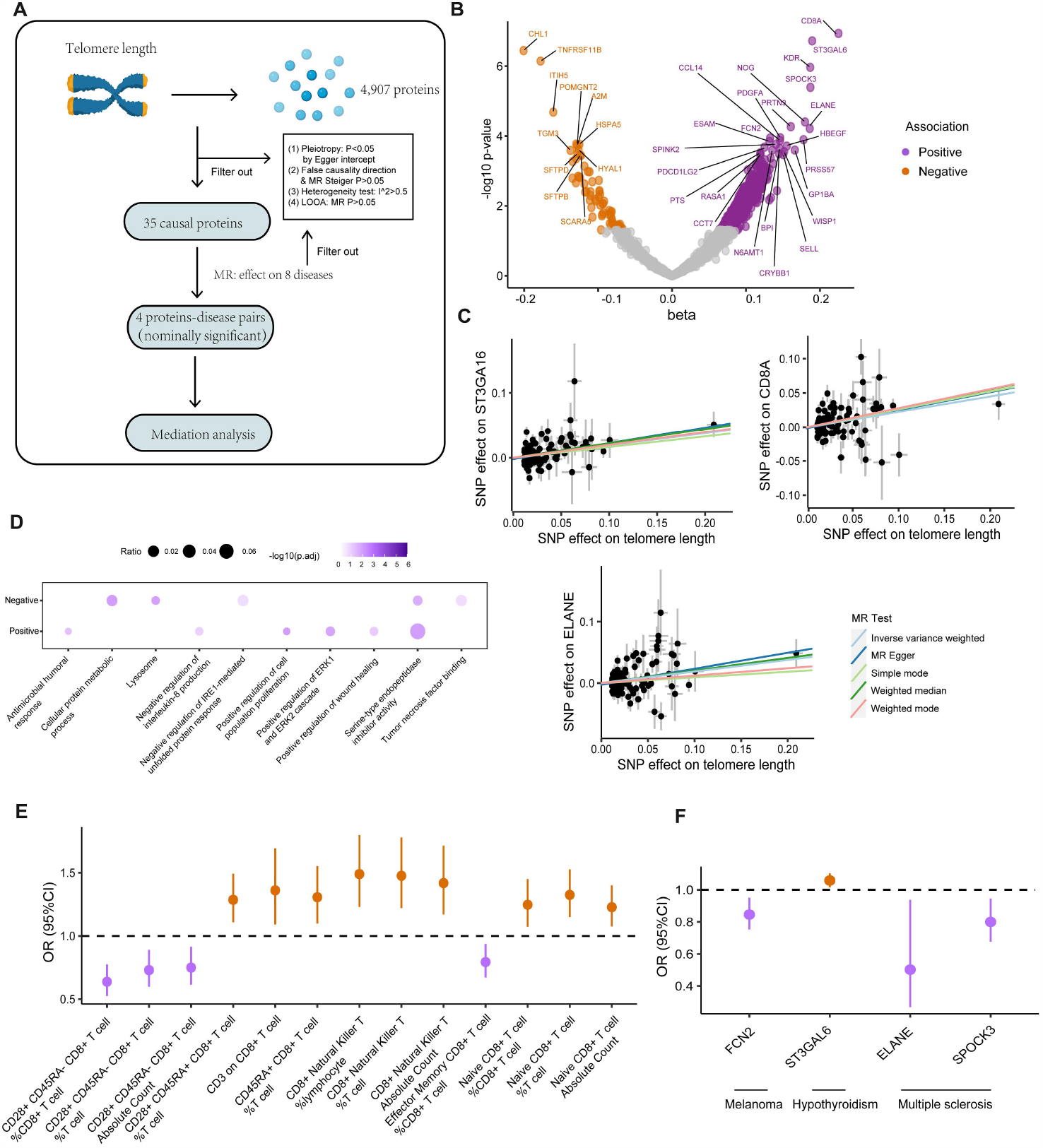
The causal effect of TL on proteins. (**A**) Design overview for the investigation of the causal effect of TL on proteins. FDR, false discovery rate; LOOA, leave-one-out analysis. (**B**) Volcano plots of MR results for the causal effect of TL on plasma proteins. Labeled proteins are 35 proteins examined in primary analysis with a significant FDR-corrected *p*-value<0.05. (**C**) Scatter plot for the causal effect of TL on the selected protein levels. (**D**) Enrichment analysis of biological functions for the 24 positively and 11 negatively influenced proteins by TL. (**E**) Causal effects of TL on 13 CD8+ T subpopulations. (**F**) Causal effects of the selected proteins on diseases. The dots are the causal estimates on the OR scale, and the whiskers represent the 95% confidence intervals for these ORs. OR, odd ratios. TL, telomere length.

### Causal effect of TL-driven proteins on 8 diseases

To test the second hypothesis, that proteins mediate the impact of TL on diseases, a two-sample MR was initially performed to evaluate the causal effect of 35 TL-driven proteins on 8 diseases. Following the *cis*-pQTL search of 35 proteins and data harmonization, 27 proteins were examined in the MR analysis. After MR analysis and procedure filtering, no statistical significance was observed for 27 proteins on diseases on a level of Benjamini-Hochberg multiple testing correction. However, 4 associations were discovered at the nominally significant (uncorrected *p*<0.05) level, as validated by sensitivity and leave-one-out analysis (**Figure 2F**). It was discovered that increasing blood ST3GAL6 expression by one SD was related to an increased incidence of hypothyroidism (OR: 1.058; 95%CI: 1.015-1.103; *p*=0.0076). Per 1-SD rise in blood SPOCK3 and ELANE expression, the incidence of multiple sclerosis was reduced, with ORs (95%CI) of 0.79 (95%CI: 0.67-0.94; *p*=0.0092) and 0.502 (95%CI: 0.268-0.938, *p*=0.030), respectively. Furthermore, each 1-SD increase in blood ST3GAL6 expression was linked to a lower incidence of hypothyroidism (OR: 1.058; 95%CI: 1.015-1.103; *p*=0.0075). However, there was no evidence that these 4 TL-driven proteins mediated the effect of TL on individual diseases due to the opposite direction of the total effect (β_TL-to-disease_) with the mediated effect (β_TL-to-protein_×β_protein-to-disease_) (**Supplemental Table 1**).

Overall, we conducted a large-scale MR investigation that sought underlying mechanisms that link plasma proteins, TL, and 8 diseases (**Figure 3**). PSMB4, PARP1, GDI2, MAX, GMPR2, ARPC1B, ATOX1, and NUDT5 were found to be genetically linked to TL. Of interest, 21 pairs have revealed that the length of telomeres mediates the causative effect of circulating proteins on diseases. These findings provide insights into telomere biology and prioritize the identified proteins as potential intervention targets for the disease. Future research with a larger and more distinct genetic background should be used to validate these results.

**Figure 3.**
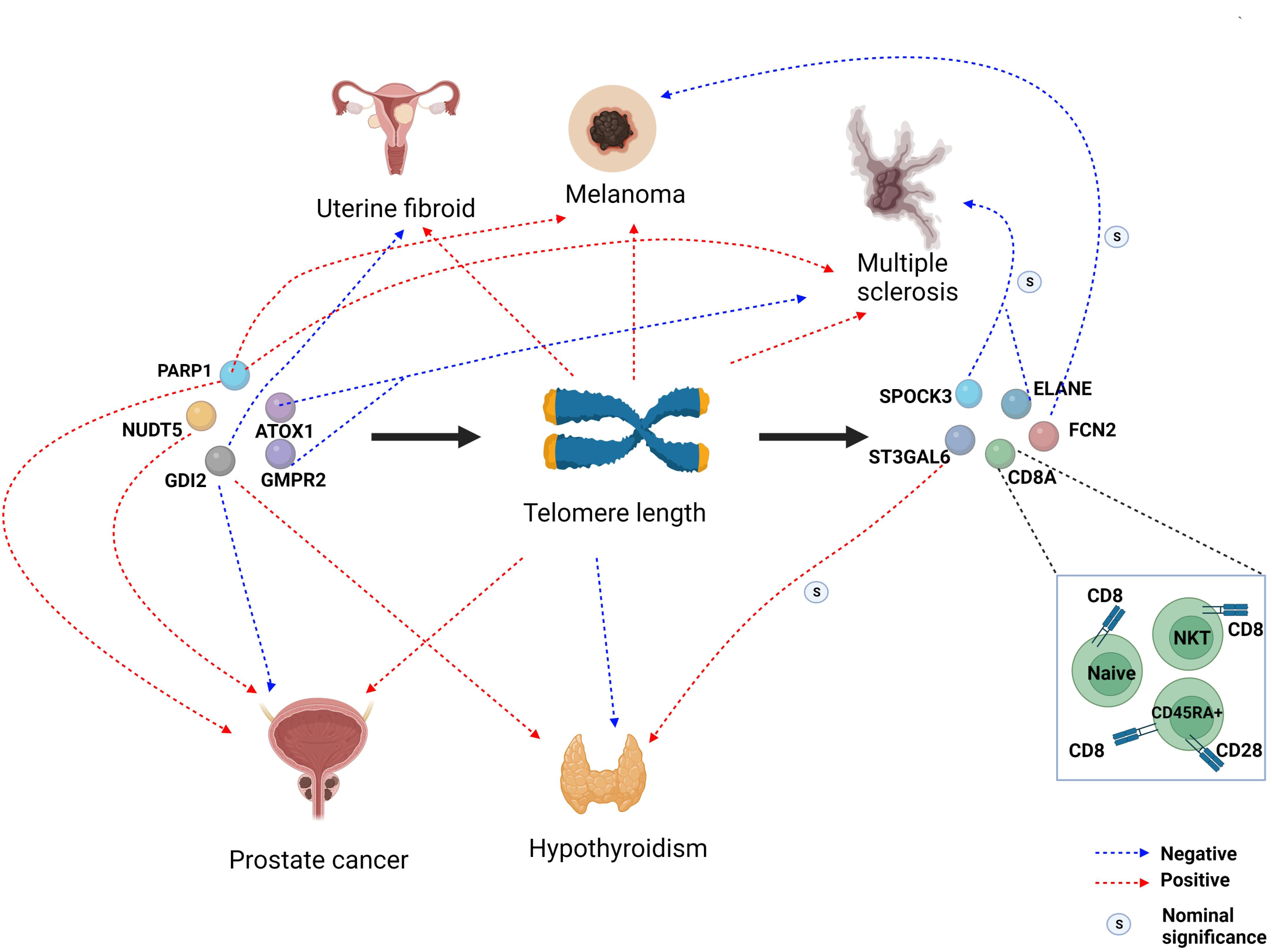
Schematic representation of the results we obtained in the study.

## Supporting information

Supplementary Table 1. The detailed GWAS information used in our investigation, as well as the summary statistics obtained in this study.

## Data Availability

All data produced in the present work are contained in the manuscript

## Acknowledgments

The authors thank all of the researchers from the related genetic consortia and GWAS for making the data available to the public.

## Contributions

SF: study conception and design, data curation and analysis, and writing and reviewing of the manuscript. MJ: data curation, as well as manuscript preparation and review.

## Figure Legends

**Figure S1.**
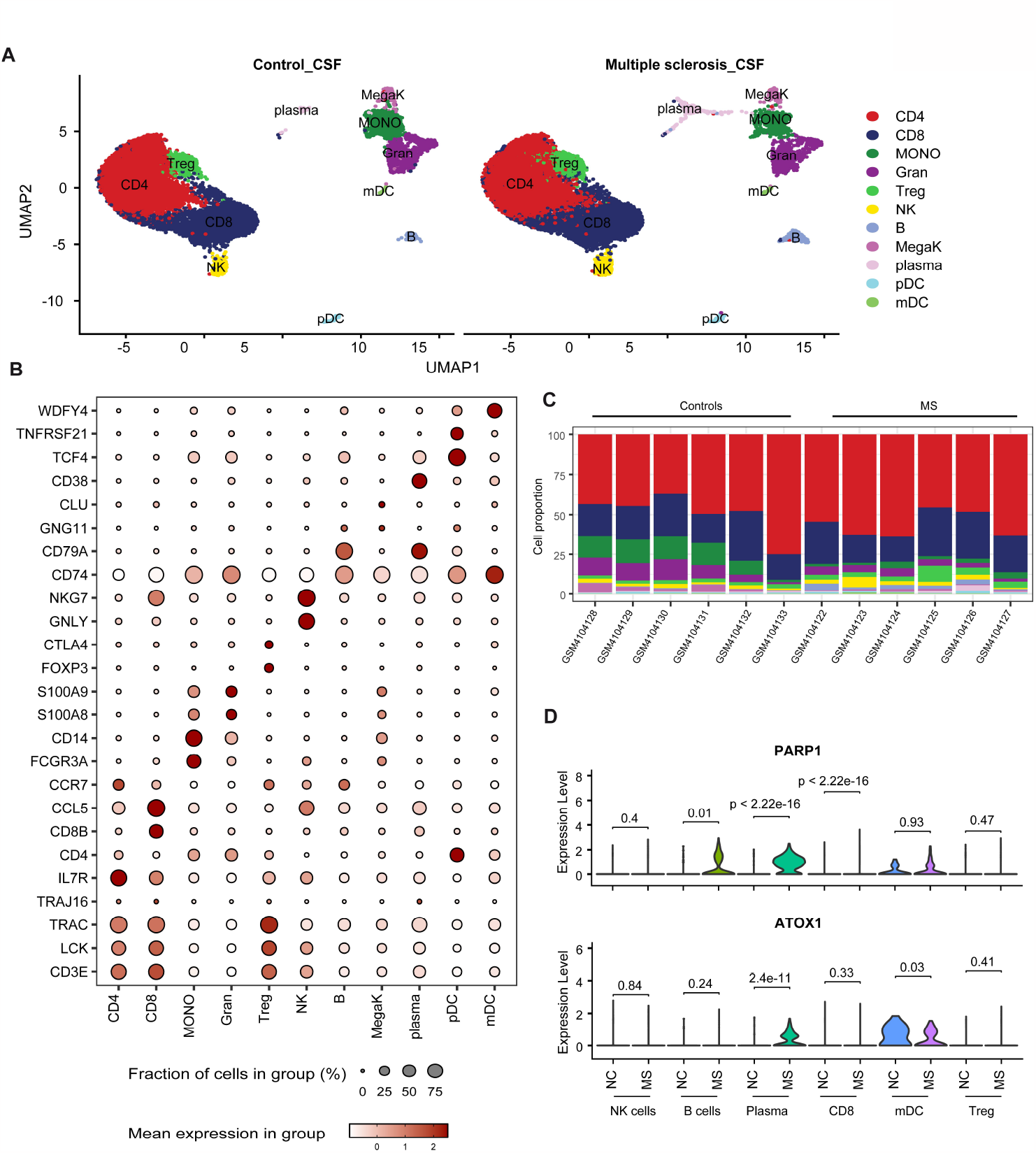
Single-cell transcriptomic atlas of cell types in the cerebrospinal fluid of multiple sclerosis patients and controls. (**A**) UMAP visualization of 33,797 cells in cerebrospinal fluid from controls (n=6) and patients with multiple sclerosis (n=6). (**B**) A dot-plot depicting the expression of marker genes in each cell type. (**C**) Barplots showing the proportion of cell types in each sample. (**D**) The differential expression levels of PARP1 and ATOX1 between controls and MS samples. CSF, cerebrospinal fluid; MS, multiple sclerosis.

**Supplementary Table 1. The detailed GWAS information used in our investigation, as well as the summary statistics obtained in this study**.

